# Outcomes of non-hospitalized isolation service during COVID-19 pandemic

**DOI:** 10.1101/2023.04.19.23288791

**Authors:** Amarit Tansawet, Pongsakorn Atiksawedparit, Pongsathorn Piebpien, Patratorn Kunakorntham, Unyaporn Suthutvoravut, Vanlapa Arnuntasupakul, Pawin Numthavaj, Atiporn Ingsathit, Oraluck Pattanaprateep, Ammarin Thakkinstian

## Abstract

**Background:** Severe acute respiratory syndrome coronavirus 2 (SARS-CoV-2) infection or COVID-19 affected more than 500 million patients worldwide and overwhelmed hospital resources. Rapid increase of new cases forced patient isolation to be conduct outside the hospital where many strategies have been implemented. This study aimed to compare outcomes among non-hospitalized isolation service.

**Methods:** A retrospective cohort study was conducted in asymptomatic and mildly symptomatic adult patients who were allocated to home isolation, community isolation, and hospitel (i.e., hotel isolation) under service of Ramathibodi Hospital and Chakri Naruebodindra Medical Institute. Variables including patients’ characteristics, comorbidities, symptoms, and medication were retrieved for use in inverse-probability-weighted regression adjustment model. Risks and risk differences (RDs) of death, oxygen requirement, and hospitalization were estimated from the model afterward.

**Results:** A total of 3869 patients were included in the analysis. Mean age was 41.8 ± 16.5 years. Cough was presented in 62.2% of patients, followed by hyposmia (43.7%), runny nose (43.5%), sore throat (42.2%), and fever (38.6%). Among the isolation strategies, hospitel yielded the lowest risks of death (0.3%), oxygen requirement (4.5%), and hospitalization (3.3%). Hospitel had significantly lower oxygen requirements and hospitalization rates compared with home isolation with the RDs (95% CI) of -0.016 (−0.029, -0.002) and -0.025 (−0.038, -0.012), respectively. Death rates did not differ among isolation strategies.

**Conclusion:** Non-hospitalized isolation is feasible and could ameliorate hospital demands. Given the lowest risks of death, hospitalization, and oxygen requirement, hospitel might be the best isolation strategy.

## Introduction

COVID-19 is infection of the severe acute respiratory syndrome coronavirus 2 (SARS-CoV-2) which mainly affects to respiratory system. Severity of symptoms ranged from asymptomatic, running nose, cough to respiratory failure. Pneumonia could develop in 5-10% of the cases.^1^ Currently, the pandemic of COVID-19 affected more than five hundred million patients worldwide with approximately 6% death rate. ^2, 3^ Since 2020, there were approximately 4.5 million confirm cases in Thailand with 30,000 deaths. In 2021, the incidence obviously increased during July 2021 to November 2021 with 5000 to 22,000 cases/day. Moreover, rapid increase of incidences was observed in early 2022 with incidence 28,000 cases/day. However, these rates subsequently declined in May 2022.^2, 4^ COVID-19 pandemic do not affect only individual patient, it also overwhelmed hospital resources (e.g., hospital staff, medication and medical equipment, etcetera). And this leads to delayed treatment of other diseased which required hospitalization.

Apart from medication and medical equipment and based on limited evidences, isolating of confirmed cases from the others was the initially effective recommendation to control the pandemic.^5-7^ In Thailand, hospitalization of all confirmed cases has been implemented since the first confirmed case was found.^8^ However, the vast influx of confirmed cases during early 2020 overwhelmed hospital resources. And this led to, very insufficient hospital space and man power (i.e., health care provider and non-health care provider). To reserve hospital facilities, several strategies, including prioritizing patient based on severity and allocating patient to appropriate level of care, has been deployed. ^9-11^

Once COVID-19 is confirmed, clinical symptoms and risk factors of pneumonia were used to prioritize patients into mild (green), moderate (yellow) and severe (red) groups. Only red group were initially hospitalized, whereas green and yellow are allocated to isolation at home (home isolation), at public shelter (community isolation), or modified hotel (hospitel).^12^ Thus, non-hospitalized isolation service was crucial for ameliorating hospitalization and sparing hospital beds for severe cases.

Currently, 3%-6% hospital admission after home monitoring program were reported. ^13-15^ Ramathibodi Hospital and Chakri Naruebodindra Medical Institute, medical schools under Mahidol University, established non-hospitalized services targeting asymptomatic or mildly symptomatic COVID-19 patients. The non-hospitalized services included hospitel (i.e., the temporary hospital set up in the hotel), home isolation, and community isolation. Although, there were evidences determine safe, decreasing admission rate and cost reduction of hotel and home-based isolation, however direct comparison their efficiencies are limited.^13, 16, 17^ Therefore, this study aimed to compare the outcomes following non-hospitalized services regarding oxygen requirement, hospitalization, and death among 3 strategies.

## Material and methods

Data of COVID-19 patients treated from April to November 2021 at Ramathibodi Hospital and Chakri Naruebodindra Medical Institute were retrieved for analysis after approval of the ethics committee. Patients would be eligible if asymptomatic or mildly symptomatic and aged 15 years or older. Only data from patients with a complete record of symptoms and comorbidities required for adjustment in the treatment effect (TE) model, were used. The outcomes of interest included hospitalization, oxygen requirement, and death.

### Triage and Service types

After Covid-19 infection is confirmed, clinical symptoms and risk factors of pneumonia are assessed by hospital staffs via telephone. Triage level and their criteria was described in Table 1.^18^ Green and yellow groups were assigned to non-hospitalized service which was divided into three types (i.e., home isolation, community isolation, and hospitel) depended on patient’ family or community supports availability. During isolation, clinical monitoring, communicating with health care providers, and treatments (i.e., antiviral agents, supportive medications, and oxygen therapy) were delivered. Details of each service are shown in Table 2. Twenty-four hours infectious disease specialists’ consultation were also available for all type of isolation; but routine infectious disease specialist’s round was provided only in hospitel. Chest X-ray was not available for home isolation.

**Table 1.** Triage categories and their criteria.

| Colors | Criteria |
| --- | --- |
| Green | Asymptomatic or mild symptoms (i.e., fever, cough, running nose, red eye, anosmia, sore throat, rash, diarrhea) |
| Yellow | Mild symptom with risk factors of severe COVID-19 as the following:<br>1. > 60 years old<br>2. Chronic lung diseases (e.g., COPD and asthma)<br>3. Chronic kidney disease<br>4. Cardiovascular disease<br>5. Diabetic mellitus<br>6. Cirrhosis<br>7. Obesity (body weight > 90 kilograms)<br>8. Immunocompromised hose |
| Red | Severe symptom including:<br>1. Dyspnea or dyspnea on exertion<br>2. Evidence of pneumonia<br>3. SPO2 < 96%<br>4. Exercise induced hypoxemia |

**Table 2.**
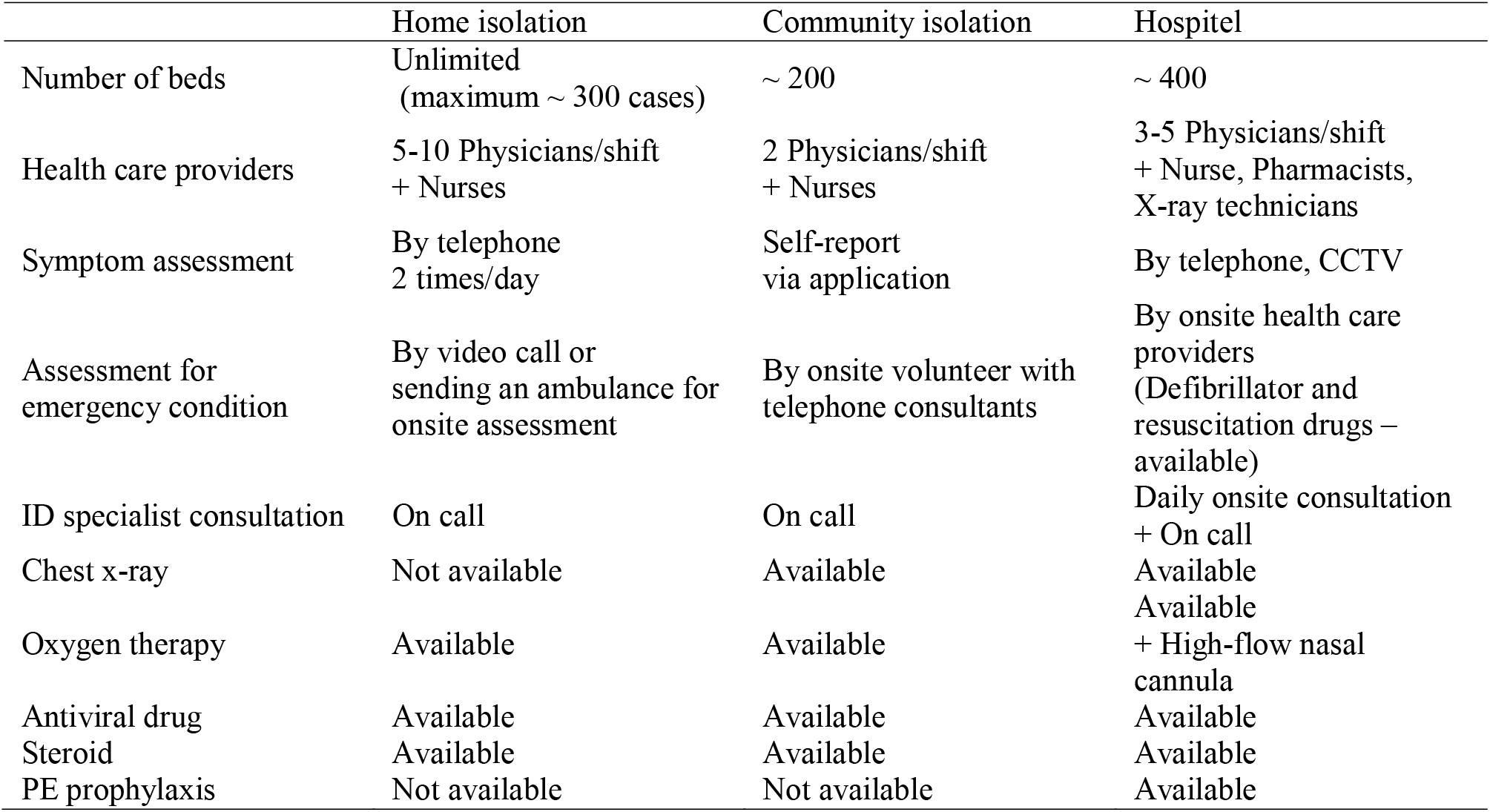
Characteristics of non-hospitalized isolation and care service

### Statistical analysis

Baseline characteristics, symptoms, and antiviral medications were described by frequency and percentage except for age which was described by the mean and standard deviation (SD). Categorical data were compared among service types using chi-square or Fisher’s exact test. Analysis of variance (ANOVA) was used for age comparison.

This study used the TE model with inverse-probability-weighted regression adjustment (IPWRA) to estimate effect size. This TE-IPWRA model consisted of two sub-models: the treatment (i.e., isolation types) assignment model applying multinomial logistic regression and the outcome model applying logistic regression. The inverse probability of being in the observed care service, obtained from the treatment assignment model, was a weight further applied in the outcome model. For covariates selection in each model (i.e., treatment assignment and outcome models), univariate analysis was performed first, followed by a multivariate model with backward elimination. The treatment (i.e., isolation types) assignment model retained only covariates significantly associated with service type allocation and the outcomes. Likewise, only covariates significantly associated with the outcome remained in the outcome model. Subsequently, the risk of the outcome occurrence (or potential-outcome mean) and risk difference (RD, or average treatment effect) between service types was estimated from the TE-IPWRA model for each outcome of interest. The conditional independence and overlap assumptions, needed for treatment effect estimation, were checked by demonstrating covariate balance among isolation strategies and adequate overlap of the probability of being assigned to each service, respectively. STATA 17 was the statistical program used throughout the analysis. A p-value < 0.05 was considered significance.

## Results

From 7478 patients, data were electronically available for analysis using TE-IPWRA in 3869 patients. The mean age (SD) was 41.8 (16.5) years. Forty-four percent of patients were male. Obesity, chronic obstructive pulmonary diseases, and cardiovascular diseases were observed in 7.6%, 2.9%, and 19.9%, respectively. Among isolation types, most of the comorbidities did not differ. Cough was presented in 62.2% of patients, followed by hyposmia (43.7%), runny nose (43.5%), sore throat (42.2%), and fever (38.6%). All symptoms, but diarrhea, significantly differed among service types, see Table 3. Apart from other supportive medicines, favipiravir was the most used medication in each service: 31.2%, 41.0%, and 45.5% in home isolation, community isolation, and hospital, respectively. Andrographolide, a Thai herbal medicine, was used in 13.3% of patients. Few patients (2.7%) received both favipiravir and andrographolide before this combination was abandoned due to an increase in liver toxicity. Among isolation types, hospitel yielded the lowest risks of death (0.3%), oxygen requirement (4.5%), and hospitalization (3.3%), see Figure 1. The highest risks of death and oxygen requirement belonged to community isolation, corresponding with the risks of 0.6% and 6.5%, respectively. The highest risk of hospitalization (5.8%) was observed in home isolation.

**Table 3.**
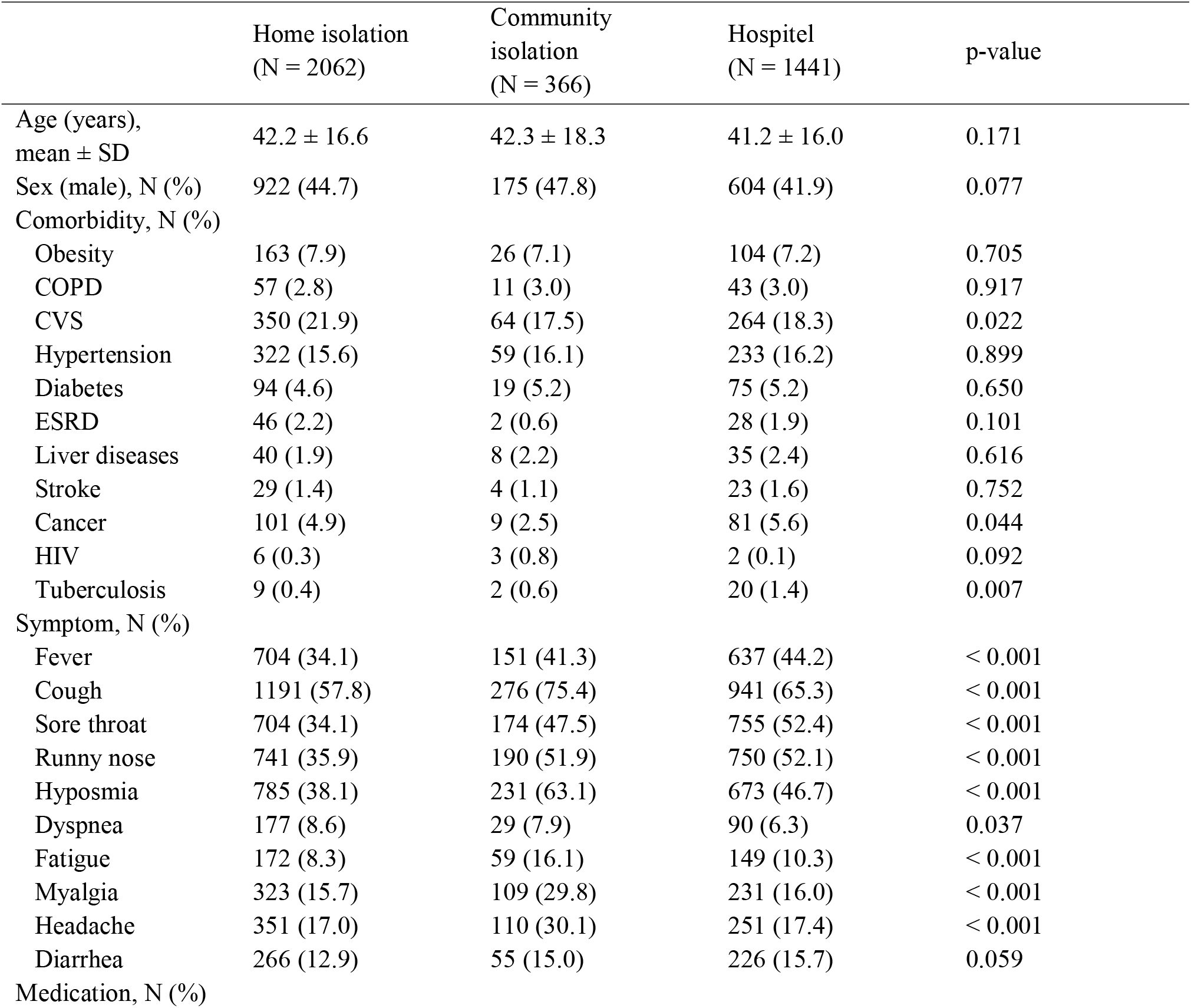
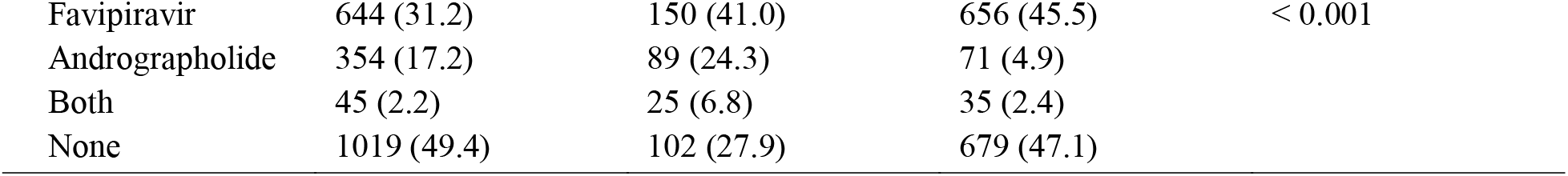
Baseline characteristics, COVID-19 symptoms, and medication used

**Figure 1.**
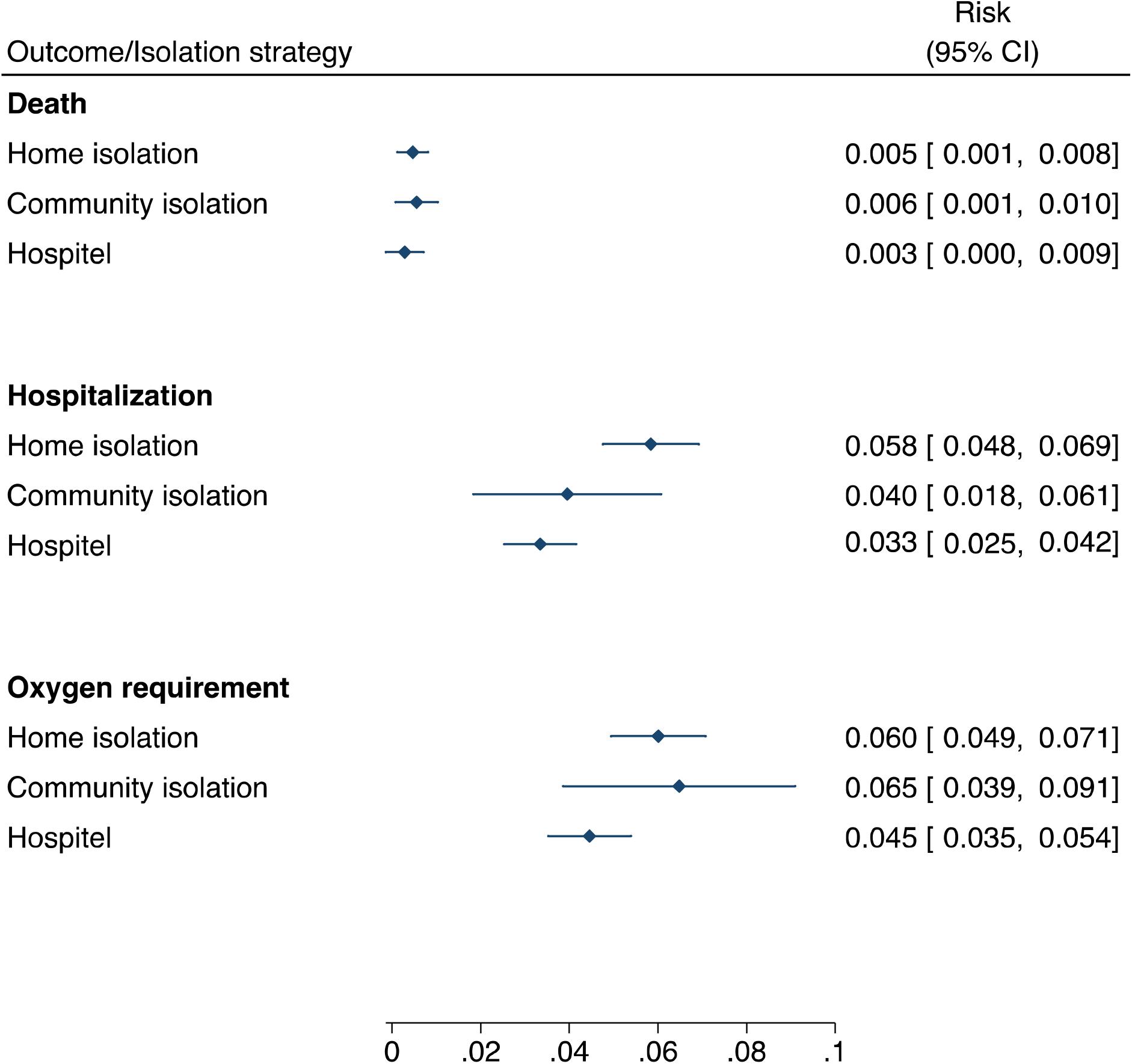
Risk of each outcome regarding isolation strategy

Among comparison, only hospitel had significantly lower oxygen requirements and hospitalization risks compared with home isolation. The RDs (95% CI) were -0.016 (−0.029, -0.002) and -0.025 (−0.038, -0.012), respectively, see Table 4. In other words, hospitel has sixteen fewer oxygen requirements and twenty-five fewer hospitalizations for every 1000 patients than home isolation. Even though hospitel has twenty fewer oxygen requirements and six fewer hospitalizations for every 1000 patients than community isolation, the difference was not statistically significant. Two and three fewer deaths were observed for every 1000 patients admitted to hospitel compared with home and community isolation. However, the risk of death did not significantly differ in every comparison. No significant difference was observed in comparing community versus home isolation.

**Table 4.** Risk difference of each outcome among isolation strategies

| Comparison | RD (95% CI) |  |  |
| --- | --- | --- | --- |
|  | Hospitalization | Oxygen requirements | Death |
| Hospital vs | -0.025 | -0.016 | -0.002 |
| Home isolation | (-0.038, -0.012) | (-0.029, -0.002) | (-0.008, 0.005) |
| Community isolation vs | -0.019 | 0.005 | 0.001 |
| Home isolation | (-0.042, 0.005) | (-0.023, 0.033) | (-0.005, 0.007) |
| Hospital vs | -0.006 | -0.020 | -0.003 |
| Community isolation | (-0.029, 0.016) | (-0.048, 0.007) | (-0.010, 0.005) |

## Discussion

This study indicated that for asymptomatic and mildly symptomatic COVID-19 patients, hospitel was the best strategy to avoid oxygen requirement and death, followed by home and community isolation. Again, hospitel is the best considering hospitalization. Community isolation was better than home isolation in hospitalization avoidance. However, this study failed to demonstrate a significant difference in most comparisons.

With the rapid increase of new cases, the healthcare system would be collapsed, and less formal systems set up outside the hospital were inevitable. Isolation, combined with quarantine, contact tracing, and large-scale screening, has been proven effective in disease spreading control.^5-7^ In addition, isolation, integrated with non-hospitalized care service, could ameliorate hospital demand.

In the early days of the outbreak, hospitel was set up for asymptomatic patients, while mildly and severely symptomatic were hospitalized. But as the number of critically ill patients increased, hospitel was subsequently adapted to be able to care for both asymptomatic and mildly symptomatic cases. At the peak of the outbreak, even hospitel was insufficient, and other isolation strategies (i.e., home and community isolation) were later deployed.

Home isolation depended on telemedicine and logistics. The advantage of home isolation is cost-saving and better quality of life.^19, 20^ This strategy is perfect for patients whose household infrastructure is not a problem. However, an overcrowded community where many people live together in a small accommodation required another solution^21^ that was either hospitel or community isolation. Community isolation was set up using public places in those communities and operated by medical personnel and volunteers. Both home and community isolation strategies were intended for treating asymptomatic and mildly symptomatic patients. Unfortunately, many severe cases were unavoidably treated during the outbreak’s peak.

Even though death was not statistically differed among isolation strategies, 0.3%, 0.5 %, and 0.6% death rates were observed from the patients initially triaged as asymptomatic or mildly symptomatic and allocated to the hospitel, home isolation, and community isolation, respectively. These findings might reflect the burden of pandemics, which all resources were consumed, including man powers, hospital beds, and medication supplies. During the peak of outbreak, patient monitoring might be far from flawlessness. Due to its fixed capacity, hospitel could achieved a closer and more effective monitoring than the others. Therefore, its death rate was the lowest.

The highest death and oxygen requirement in the community isolation group might be explained by difference care process and patient assessment which possibly resulted in delayed detection of severe disease. In community isolation, the care process was mostly operated by volunteers, and the patients’ self-report via mobile phone application was a primary route of daily assessment. These results also suggested that criteria used for allocation to different isolation strategies might require revision. Perhaps, only asymptomatic cases should be allocated to the community isolation.

Hospitalization, ranged from 3.3% to 5.8 %, did not contradict other studies.^13-15^ The highest hospitalization rate was observed in the home isolation patients. The hypothesis is that the patients isolated at their home might feel insecure and require more medical attention when their symptoms altered.

The ideal study design for treatment strategy evaluation should be a randomized control trial (RCT). However, numerous factors could intervene during the pandemic, and conducting RCT is a challenging task. This study used real-world data with the appropriate statistical model to emulate RCT. By balancing covariates among service allocation, results should be valid.

Nevertheless, this study has some limitations. First, a chest x-ray was not performed on every asymptomatic or mildly symptomatic patient, especially in the home isolation group, which made ascertainment of pneumonia uncertain. Hence, this study did not investigate pneumonia outcomes. Second, some asymptomatic or mildly symptomatic patients progressed to severe COVID-19 pneumonia, or their comorbidities worsened during isolation. However, all facilities were fully occupied during the outbreak’s peak, precluding patients from oxygen supplements and hospitalization. Oxygen requirements and hospitalization rates might be spurious low. Even that death rate should reflect the actual disease burden among isolation strategies. Third, the number of patients in the community isolation was relatively low; this group’s estimate was less precise. Low precision might explain why significant results were not reached in comparing hospitel versus community isolation. Finally, this study did not examine the cost of each isolation strategy. No information was provided on whether hospitel was more cost-effective than the others. Economic evaluation should be further conducted.

In conclusion, hospitel resulted in the lowest risk of hospitalization, oxygen requirements, and death. Hospitel had a significantly lower risk of hospitalization and oxygen requirements than home isolation but not community isolation. No difference was observed comparing community and home isolation. Because a delta viral strain predominated during 2021, extrapolation from these findings for subsequent pandemics has to be done with caution. Moreover, cost-effectiveness of isolation strategies should be further investigated.

## Data Availability

All data produced in the present study are available upon reasonable request to the authors

## Notes

### Competing Interest Statement

The authors have declared no competing interest.

### Funding Statement

This study did not receive any funding

### Author Declarations

Ramathibodi Hospital Ethics Committee.

